# Radiomics-Based Early Triage of Prostate Cancer: A Multicenter Study from the CHAIMELEON Project

**DOI:** 10.1101/2025.04.29.25326685

**Authors:** Aikaterini Vraka, Manuel Marfil-Trujillo, Gloria Ribas-Despuig, Silvia Flor-Arnal, Leonor Cerdá-Alberich, Paula Jiménez-Gómez, Ana Jimenez-Pastor, Luís Martí-Bonmatí

## Abstract

Prostate cancer (PCa) is the most commonly diagnosed malignancy in men worldwide. Accurate triage of patients based on tumor aggressiveness and staging is critical for selecting appropriate management pathways. While magnetic resonance imaging (MRI) has become a mainstay in PCa diagnosis, most predictive models rely on multiparametric imaging or invasive inputs, limiting generalizability in real-world clinical settings. This study aimed to develop and validate machine learning (ML) models using radiomic features extracted from T2-weighted MRI—alone and in combination with clinical variables—to predict ISUP grade (tumor aggressiveness), lymph node involvement (cN) and distant metastasis (cM). A retrospective multicenter cohort from three European sites in the Chaimeleon project was analyzed. Radiomic features were extracted from prostate zone segmentations and lesion masks, following standardized preprocessing and ComBat harmonization. Feature selection and model optimization were performed using nested cross-validation and Bayesian tuning. Hybrid models were trained using XGBoost and interpreted with SHAP values. The ISUP model achieved an AUC of 0.66, while the cN and cM models reached AUCs of 0.77 and 0.80, respectively. The best-performing models consistently combined prostate zone radiomics with clinical features such as PSA, PIRADSv2 and ISUP grade. SHAP analysis confirmed the importance of both clinical and texture-based radiomic features, with entropy and non-uniformity measures playing central roles in all tasks. Our results demonstrate the feasibility of using T2-weighted MRI and zonal radiomics for robust prediction of aggressiveness, nodal involvement and distant metastasis in PCa. This fully automated pipeline offers an interpretable, accessible and clinically translatable tool for first-line PCa triage, with potential integration into real-world diagnostic workflows.

## Introduction

Prostate cancer (PCa) remains the most frequently diagnosed malignancy and the second leading cause of cancer-related death among men around the world [1]. Accurate risk stratification and staging are crucial for guiding treatment decisions, ranging from active surveillance to radical interventions. Traditional clinical assessments, including prostate specific antigen (PSA) levels, digital rectal examinations and biopsies are often complemented by imaging modalities to enhance diagnosis accuracy [1–4].

Magnetic Resonance Imaging (MRI) has become a cornerstone in the non-invasive evaluation of PCa, with many studies showing clear benefits of biparametric (bpMRI) instead of multiparametric (mpMRI) MRI [5–8]. T2-weighted imaging (T2w) offers superior soft tissue contrast, facilitating detailed visualization of prostate anatomy and lesion localization. While diffusion-weighted imaging (DWI) and dynamic contrast-enhanced (DCE) imaging provide additional functional information, T2w sequences are widely available and consistently utilized across clinical settings [6].

Radiomics, the extraction of high-dimensional quantitative features from medical images, combined with machine learning (ML) techniques, has shown promise in capturing tumor heterogeneity and improving predictive modeling in oncology [9]. In PCa, radiomics have been applied to predict tumor aggressiveness, lymph node involvement and biochemical recurrence [10–15].

Building on these imaging capabilities, recent advances in artificial intelligence have enabled the development of predictive models that integrate radiomic features with clinical data to improve staging accuracy. It has been demonstrated that radiomics-based ML models using MRI show comparable accuracy to standard nomograms for lymph node invasion (LNI), while the use of computed tomography (CT) and positron emission tomography-CT (PET/CT) models can show high diagnostic and prognostic results [16, 17]. Other studies demonstrated that ML models integrating radiomics from T2w MRI with clinical and pathological data could predict biochemical recurrence with an area under the curve (AUC) of at least 0.84 [18, 19].

However, many existing studies have limitations, including reliance on single-center, retrospective datasets with enriched populations that may not reflect real-world disease prevalence [16, 20]. Additionally, while CT and PET/CT models are commonly used for staging, their sensitivity and specificity for detecting LNI are suboptimal, particularly in early-stage disease [21, 22]. As for distant metastasis, PET/CT still remains the reference in its detection, while recent studies report high performance using PET/MRI [23, 24]. Nevertheless, this technology shows significant limitations with respect to MRI, such as exposure to radiation, limited availability and high costs [25, 26].

In this study, we present a multi-institutional analysis involving real-world data from three European clinical sites of the Chaimeleon project. We developed and evaluated three ML models trained on radiomic features extracted from T2w MRI, guided by prostate zonal segmentation masks and combined with clinical variables. Besides the exclusive use of T2w, which are widely available and the baseline imaging modality for PCa diagnosis, we propose a single pre-processing and analysis pipeline yielding the assessment of three key biomarkers for a first-level PCa triage: aggressiveness, LNI and distant metastasis. In this way, our approach aims to provide generalizable and clinically translatable tools for the stratification and staging of PCa risk.

## Materials and Methods

### Materials

Retrospective data from three European clinical sites from the Chaimeleon project were used [27]. These sites were Valencia (Spain), Porto (Portugal) and Angers (France). Due to the retrospective and de-identified use of the data, informed consent was waived. Originally, data from 534 patients with newly diagnosed PCa were recruited.

For each patient, an imaging study consisting of bpMRI of the prostate or pelvic area was included, accompanied by clinical information derived from the electronic case report form (eCRF), including the age at diagnosis, total PSA, PIRADSv2 and the three variables that would be used as ground truth during the training of our three AI models: ISUP (after histopathological confirmation), LNI-clinically confirmed (cN) and distant metastasis after clinical confirmation (cM). Information on database size and prevalence as well as clinical information for each model can be seen in Table 1. Age appears in mean and standard deviation (sd), while PSA appears in median and interquartile range (IQR) after the application of Shapiro-Wilk test on both variables to check for normality [28]. The distribution per clinical center can be observed in Figure 1, where due to the lack of ground truth in some cases, the cN and cM models database appears reduced and the only clinical site is Valencia.

**Table 1.**
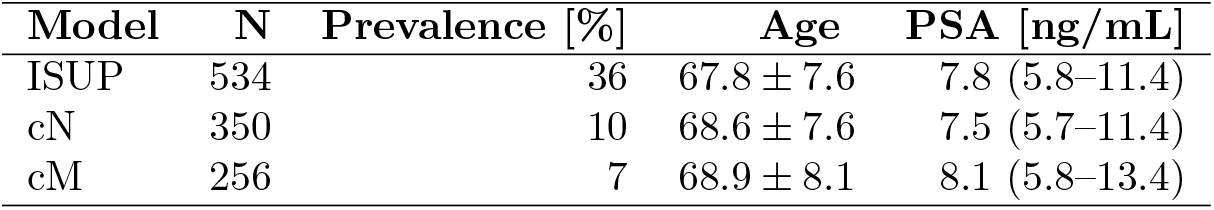
Database size (N), prevalence and summary statistics for age and PSA per model. PSA is shown in median and IQR.

**Figure 1.**
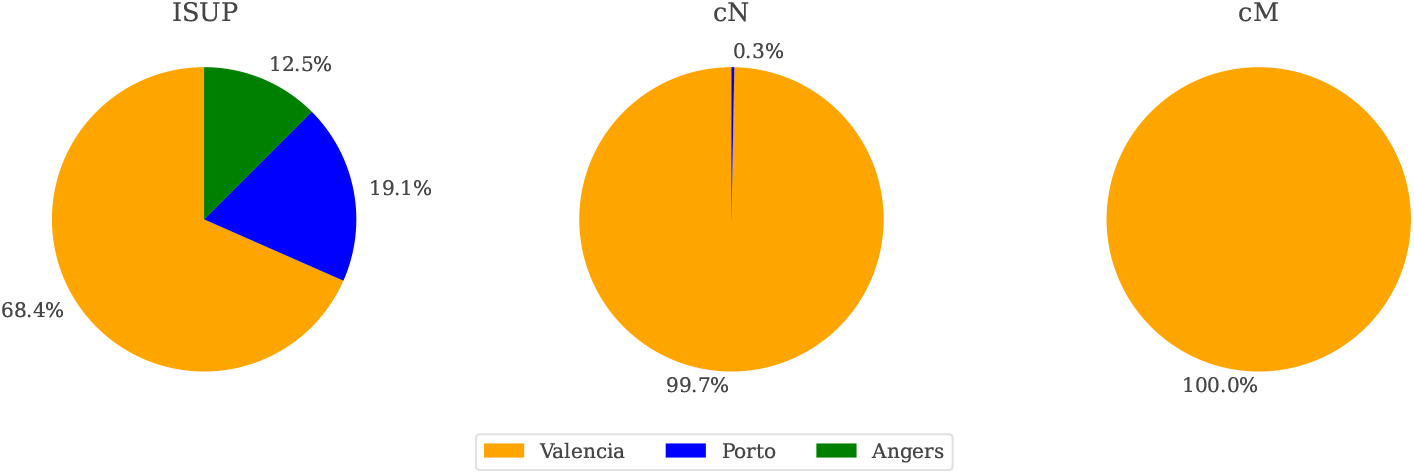
Center distribution per model. For the cN model, only one patient came from Angers, while the remaining patients came from Valencia. For the cM model, all data came from Valencia.

### Methods

Axial T2w images were selected as the only imaging input. The pre-processing and main analysis pipeline was identical for all three developments (see Figure 2), with the only difference being found in the per case selected database (rows filtering) as well as the model training process. Development was conducted in python language using Python 3.11 version.

**Figure 2.**
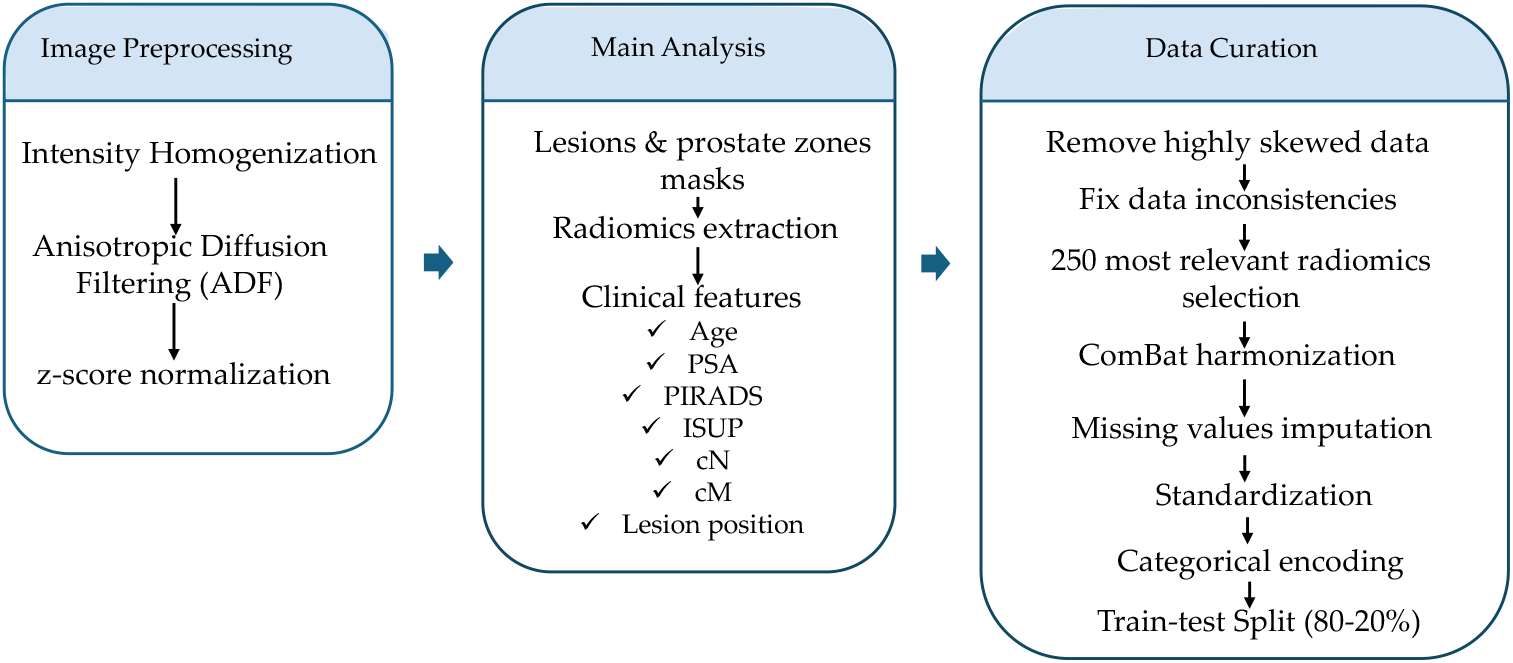
Image pre-processing, main analysis and data curation pipeline. The procedure was conducted once before the development of the three predictive models.

The first step of image pre-processing was the application of a N4 bias field correction algorithm for intensity normalization, followed by an anisotropic diffusion filter (ADF) to reduce noise while preserving prostate gland boundaries [29–31]. The last step of image pre-processing was the z-score normalization to account for differences in acquisition protocols [32]. An example of the image pre-processing result can be observed in Figure 3.

**Figure 3.**
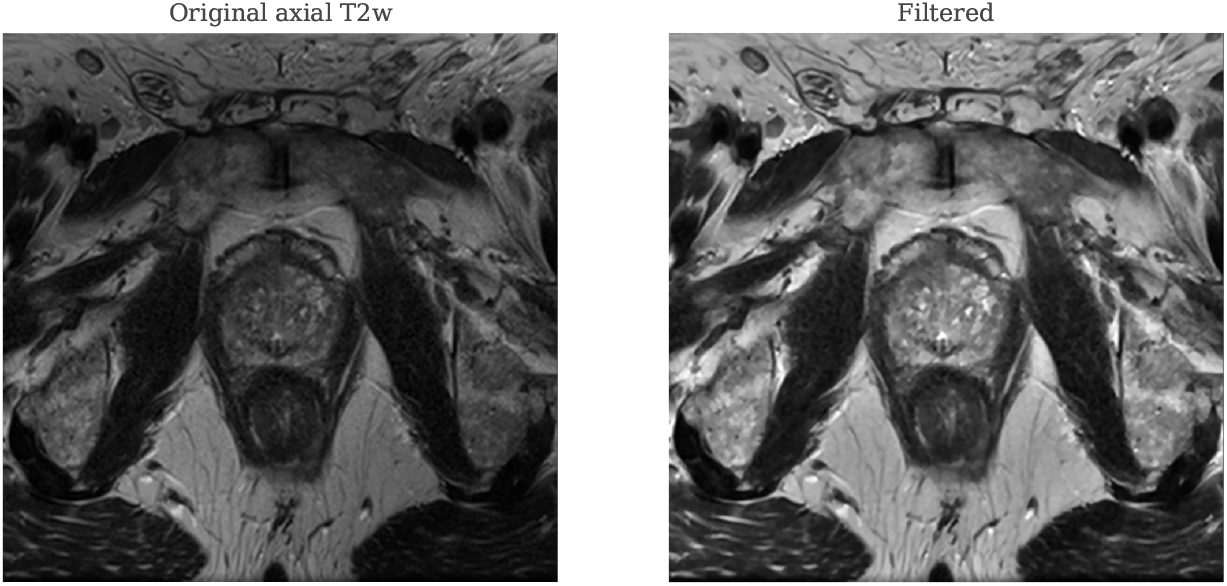
One slice of original (left) and pre-processed image (right) of a patient. Pre-processing aimed to smooth out signal intensity, preserve prostate boundaries and account for differences in acquisition protocols that were present due to the use of a multi-center database.

Segmentation masks of PCa lesions and prostate zones were acquired through QP-Prostate (Quibim, SL), as is shown in Figure 4. Radiomics were extracted using PyRadiomics with a variety of image filters. Clinical features were added to the radiomics signatures. For the prostate aggressiveness model, age, PSA and PIRADSv2 were added. For the cN and cM model, besides the above, ISUP information was recruited. For all three models, lesion position was extracted as a binary value for peripheral (PZ), central (CZ), transitional (TZ) and seminal vesicles (SV) using the lesions masks and the information was added to the model input.

**Figure 4.**
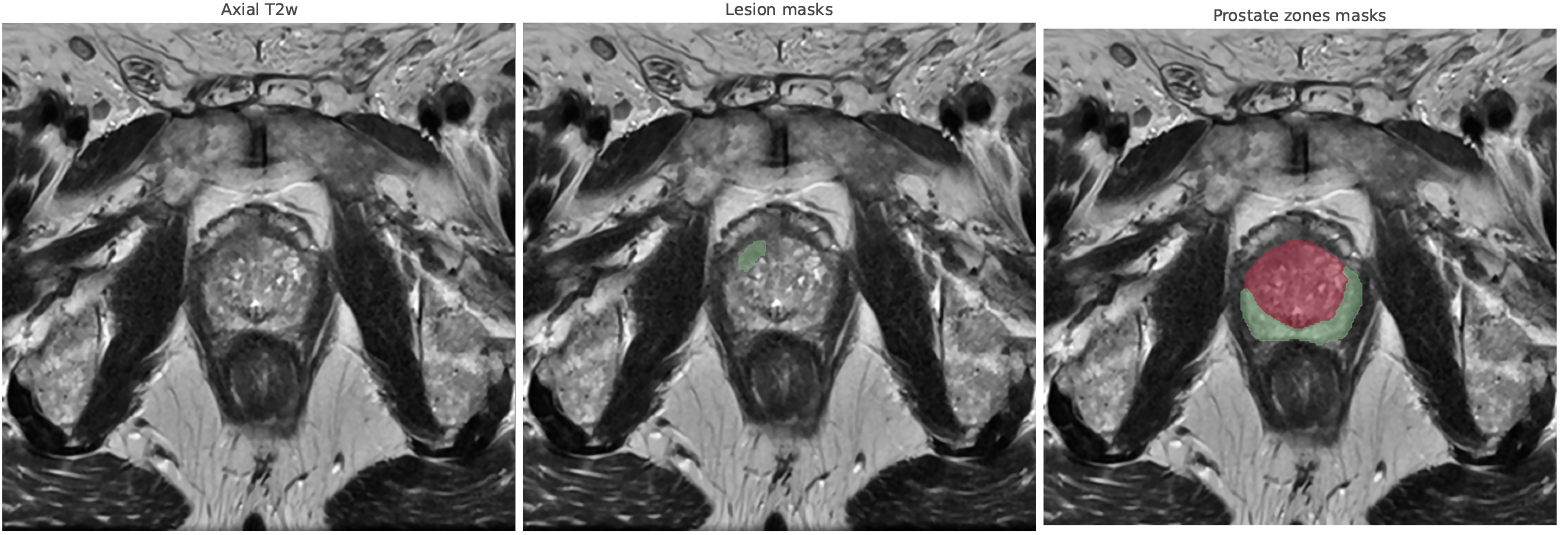
Example of a lesion and prostate zone mask for the same patient using QP-Prostate. Peripheral area is shown in green, while central and transitional area in red.

Clinical and radiomic features underwent a multi-step data curation process. Highly skewed features (skewness *>* 5.0) were removed and inconsistent or missing values were imputed using either the most frequent value (for categorical/boolean features) or K-nearest neighbors (*K* = 3) for numerical data. Boolean variables were binarized and filtered using the chi-squared test. Categorical batch-related variables (here: scanner vendor, magnetic field strength, center) were encoded and included in ComBat harmonization [33, 34]. All numerical features were standardized using z-score normalization.

Coarse feature selection was performed by applying a minimum variance threshold, removing highly correlated features (correlation *>* 0.8) and selecting the top 250 most relevant features using Minimum Redundancy Maximum Relevance (mRmR) [35]. The curated dataset was then split into training and test sets (80*/*20) using stratified sampling by target class.

Model development was carried out, for each of the three cases, using a nested cross-validation framework with five outer folds and three inner folds to ensure unbiased evaluation. Within each inner loop, mRmR feature selection was applied to identify the most relevant features, with the number of selected features ranging from 3 to 30.

Performing feature selection inside the inner loop prevents data leakage and ensures that the outer test set remains entirely independent, preserving the validity of performance estimates. To address class imbalance, the training data in each outer fold was balanced using a Synthetic Minority Oversampling Technique (SMOTE) and a range of *scale pos weight* values was also explored during hyperparameter tuning.

Separate models were trained on radiomics-only data, clinical-only data and hybrid datasets combining both. Additionally, the contribution of lesion radiomics, prostate zones radiomics and their combination was investigated.

Model optimization was performed using Optuna, a Bayesian hyperparameter search framework, with the objective of maximizing AUC on the inner folds. The best-performing feature subset and hyperparameters were then used to train an XGBoost classifier on the full training split of each outer fold. Performance was evaluated on the held-out test split (20%) using AUC, accuracy, precision, recall, F1-score and Cohen’s kappa. To enhance model interpretability, SHapley Additive exPlanations (SHAP) values were computed on the outer test predictions. SHAP values provide a unified framework for explaining individual predictions by estimating the contribution of each feature to the model’s output, enabling case-level and global interpretability of the radiomic and clinical features involved [36].

## Results

For PCa aggressiveness, a binary predictive model was trained as follows: ISUP grades*≤* 2 were considered low agrressive, while ISUP grades *>* 2 were considered highly aggressive. For cN and cM, the absence of LNI and metastasis, respectively, was considered as the positive class, while their presence was considered as the negative class.

Final models specifications are shown in Table 2. For all classification tasks, hybrid models with radiomics from prostate zones showed the highest performance [37]. For the prostate aggressiveness classification, six features were selected, three radiomics and three clinical. For the cN model, 11 features in total were selected, four of which were clinical and the remaining radiomics. For the cM model, 13 features were selected, with three being clinical and the remaining 10 radiomic signatures.

**Table 2.** Optimal configuration of the best-performing model per classification task.

| Model | Type | Radiomics | Feature number | N estimators | Max depth | Learning rate |
| --- | --- | --- | --- | --- | --- | --- |
| ISUP | hybrid | prostate zones | 6 | 58 | 3 | 0.040 |
| cN | hybrid | prostate zones | 11 | 74 | 9 | 0.025 |
| cM | hybrid | prostate zones | 13 | 233 | 5 | 0.170 |

The selected features per task are shown in Table 3. It can be observed that texture-based radiomics, such as entropy and non-uniformity as well as first-order statistics play a prominent role in all three classification tasks. PIRADSv2 and zonal location (PZ, TZ) appear consistently regardless of the task, confirming the importance of anatomical and expert-derived clinical variables. The distant metastasis (cM) model was notably more based on high-order texture characteristics, possibly reflecting increased heterogeneity in advanced disease.

**Table 3.** Selected features per classification task. Radiomic feature types are abbreviated as: GLCM: Gray-Level Co-occurrence Matrix; GLSZM: Gray-Level Size Zone Matrix; GLDM: Gray-Level Dependence Matrix; GLRLM: Gray-Level Run Length Matrix.

| PCa aggressiveness | cN | cM |
| --- | --- | --- |
| First-order: Median | GLDM: Non-uniformity | GLCM: Correlation (LHH) |
| GLSZM: Zone Entropy | First-order: 90th Percentile | GLDM: Dependence Entropy |
| First-order: Skewness | GLSZM: Non-uniformity | First-order: Median |
| PZ (Peripheral Zone) | GLRLM: Short Run Emphasis | GLDM: Variance |
| TZ (Transition Zone) | GLSZM: Emphasis | First-order: Skewness |
| PIRADSv2 | GLSZM: Zone Entropy | GLRLM: Entropy |
| — | GLCM: Inverse Difference | First-order: Minimum |
| — | PIRADSv2 | GLCM: Difference Entropy |
| — | ISUP | GLSZM: Zone Entropy |
| — | PSA | GLCM: Correlation (LLL) |
| — | PZ (Peripheral Zone) | ISUP |
| — | — | PIRADSv2 |
| — | — | TZ (Transition Zone) |

Figure 5 shows the receiver-operating characteristics (ROC) curves at the holdout test set for each classification task. The performance metrics are then summarized in Table 4. The model trained to predict prostate cancer aggressiveness (ISUP) achieved an AUC of 0.66, a balanced accuracy of 0.64 and a sensitivity of 0.61 on the test set.

**Table 4.**
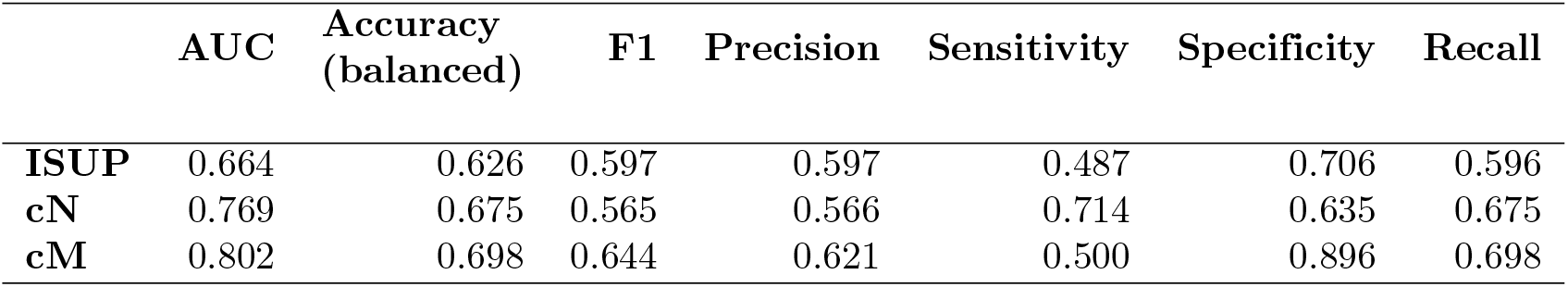
Performance matrix for the test set of each predictive task.

**Figure 5.**
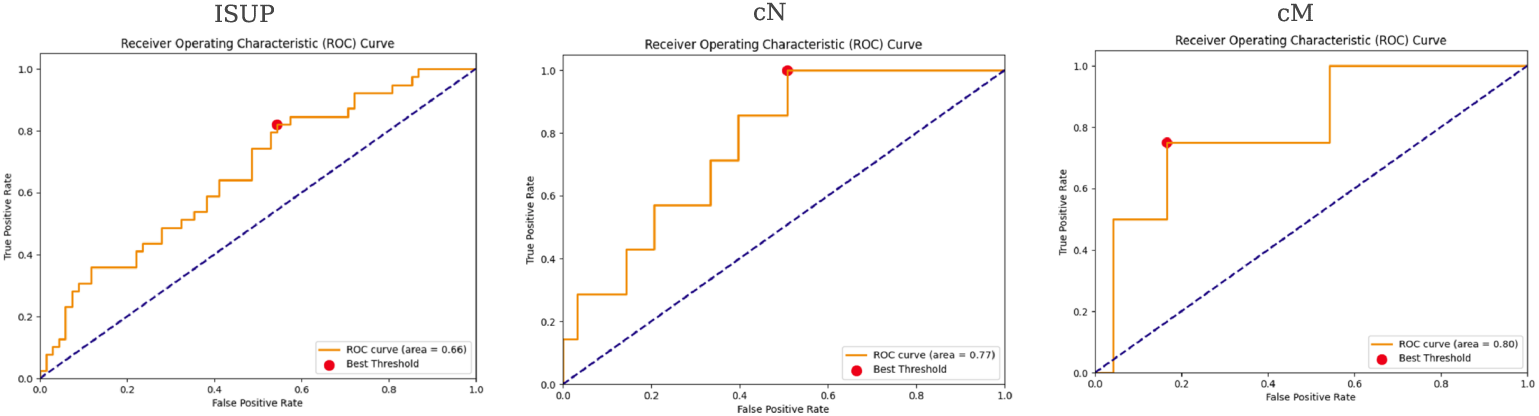
ROC curve for the test set of each predictive task.

The ROC curve reflects modest separation between aggressive (ISUP *>* 2) and non-aggressive cases. While the overall performance is moderate, the model was able to identify a substantial proportion of clinically significant tumors despite the presence of high class imbalance, possibly due to the oversampling and scaling operations during training.

The cN model achieved an AUC of 0.77, a balanced accuracy of 0.76 and a sensitivity of 0.74. Despite the low prevalence of positive LNI (10%), the model maintained strong sensitivity, which is particularly relevant from a clinical standpoint. Early and accurate detection of LNI is crucial for treatment planning and the model’s ability to identify true positives without overfitting to the dominant class reflects its robustness. The best overall performance was observed in the cM model, with an AUC of 0.80, balanced accuracy of 0.79 and sensitivity of 0.78. These values indicate strong discriminative power despite a highly imbalanced dataset (7% cases with distant metastasis). The high sensitivity is of particular clinical interest, as early detection of distant metastasis significantly influences prognosis and therapeutic decision-making. The balanced accuracy also suggests the model retained specificity while effectively detecting metastatic cases.

The confusion matrices with the true positive and true negative outcomes on the test set of each classification task can be seen in Figure 6. The ISUP model achieved a relatively balanced performance between the “Low” and “High” aggressiveness groups, though there was some degree of misclassification between the two classes. For the cN and cM models, the matrices highlighted a tendency toward higher sensitivity for the cN0 and cM0 classes, with fewer false negatives. However, specificity was lower, with some overprediction of nodal and metastatic involvement.

**Figure 6.**
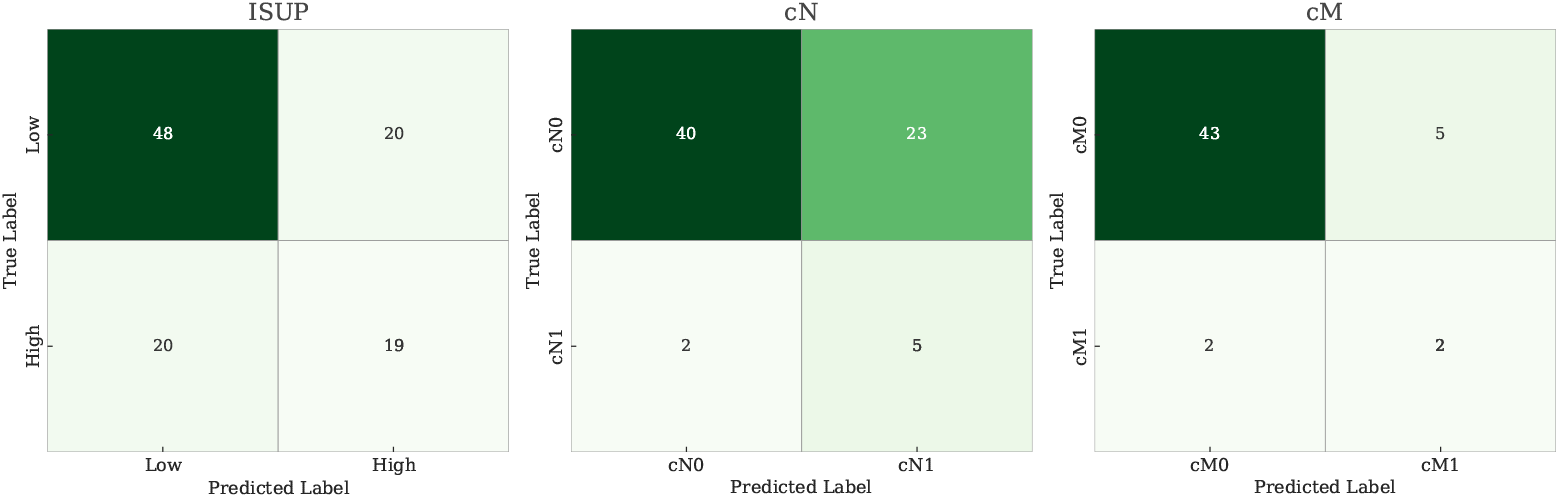
Confusion matrices for the classification of ISUP grade group (Low vs High aggressiveness), clinical LNI (cN0-no invasion vs cN1-invasion) and clinical metastatic status (cM0-no metastasis vs cM1-metastasis). True positives (TP) and true negatives (TN) are highlighted in green. The matrices show the number of correctly and incorrectly classified cases.

SHAP summary plots (Figure 7) were used to visualize the contribution of individual features to model predictions for prostate aggressiveness, cN and cM status. Across all models, both clinical variables and radiomic features from various wavelet decompositions demonstrated relevance, though the patterns of importance varied by outcome.

**Figure 7.**
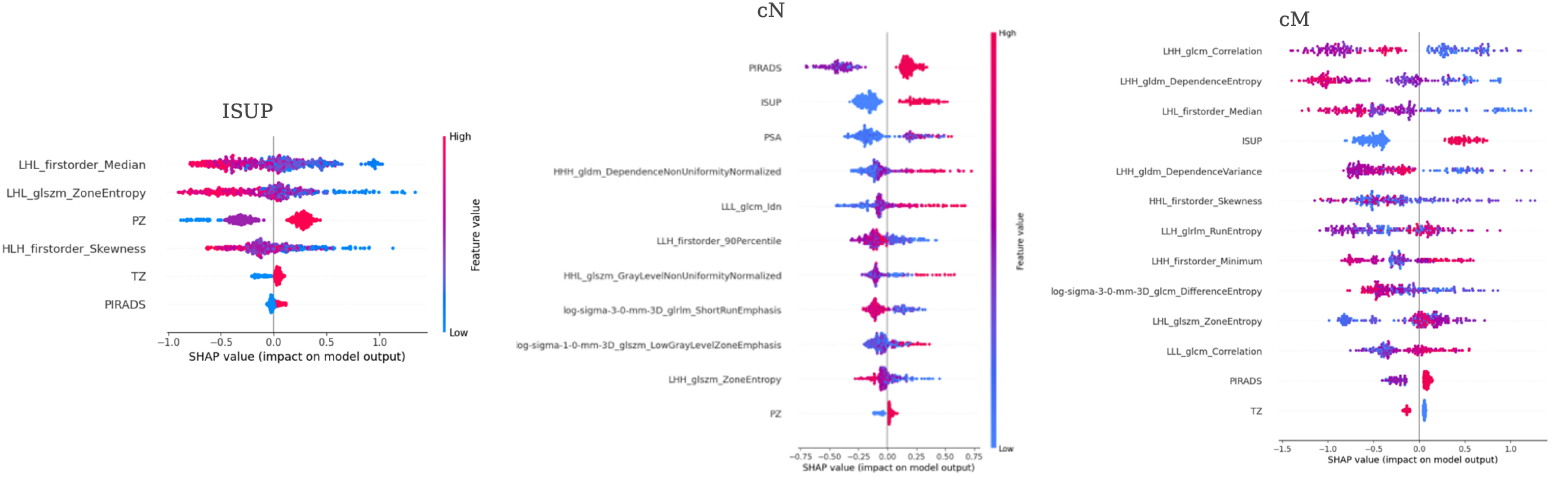
SHAP summary plots illustrating the impact of individual features on the model output for each classification task. Each point represents a patient case, colored by the value of the corresponding feature (red = high, blue = low). Features are ordered by overall importance, with SHAP values indicating the direction and magnitude of each feature’s contribution to the model’s decision.

For ISUP grade classification, the most influential features included *LHL_firstorder_Median, LHL_glszm_ZoneEntropy* and *HLH_firstorder Skewness*, suggesting that intensity and texture heterogeneity within tumor regions are associated with aggressiveness. Notably, the PZ and TZ location indicators also contributed, indicating zonal variation in predictive characteristics.

In the cN model, PIRADSv2, ISUP and PSA were among the top contributors, reinforcing their central clinical relevance. In addition, texture-based radiomic features showed consistent importance, indicating that more subtle structural tissue changes may reflect nodal involvement.

For cM status, radiomic features dominated the top rankings. Features such as *LHH_glcm_Correlation, LHH_gldm_DependenceEntropy* and *LHL_firstorder_Median* were strong contributors, while ISUP and PIRADSv2 had lower influence. This suggests that the model may be capturing latent imaging signatures of metastatic potential not directly represented in standard clinical scores.

Across all tasks, SHAP values highlighted not only the magnitude of feature impact but also the directionality: for example, higher values of certain entropy-related features were generally associated with increased predicted risk (positive SHAP values). The color gradients (blue to red) further revealed interactions between feature magnitude and effect, offering deeper insights into feature behavior across patient subgroups.

These results reinforce the utility of combining radiomics with clinical parameters to enhance model explainability and clinical insight, potentially guiding personalized risk stratification strategies.

## Discussion

In this study, we present a robust, interpretable and unified ML framework for the first-level triage of prostate cancer aggressiveness, LNI and distant metastasis using only T2w MRI and routinely available clinical data. Our results demonstrate competitive performance when compared to more complex state-of-the-art (SOTA) models that typically require multiparametric imaging, biopsy-derived data or high-cost PET/MRI.

Several recent studies have reported strong predictive performance using mpMRI or PET/MRI combined with radiomic ML models. For example, Bourbonne et al. developed a multimodal MRI-based model to predict LNI, achieving a C-index of 0.89 by integrating radiomics with clinical variables [12]. Similarly, Ayyıldız et al. trained ML models on biparametric MRI for distinguishing clinically significant PCa, reaching AUCs up to 0.86 [38]. While these values exceed our results, they rely on broader imaging modalities or curated datasets with enriched populations. Interestingly, when T2w-only images are used, an AUC of 0.64 is obtained [38].

Our pipeline, by contrast, utilizes T2w MRI alone, which is widely available and routinely acquired in prostate cancer diagnosis. This design offers significant advantages in terms of clinical scalability, lower cost and ease of integration into existing workflows, especially in settings where diffusion or contrast-enhanced sequences are unavailable. Our findings are further supported by recent work from Yi et al., who demonstrated that T2w-only meta-learning models can achieve comparable accuracy to mpMRI-based methods [39].

When comparing task-specific performance, our results for cN and cM (AUCs: 0.77 and 0.80) are particularly noteworthy given the real-world, imbalanced, multi-institutional data used for training. Recently, a study achieved an AUC of 0.90 for the prediction of distant metastasis using a semi-automated deep learning framework which integrated bpMRI, radiomics and clinical data [40]. Nevertheless, the reported prevalence (30 *−* 43%) was substantially higher than real-world rates, potentially inflating predictive performance and limiting generalizability [41]. Other studies using advanced PET/MRI pipelines report AUCs in the range of 0.90–0.94 for predicting biochemical recurrence or overall risk [42] but at the cost of accessibility and patient exposure to ionizing radiation. Our model maintains strong performance without such trade-offs.

In terms of ISUP classification, although the AUC of 0.66 is modest, it is consistent with prior findings on PIRADSv2 category 3 lesions, where radiomic models achieve AUCs around 0.64–0.68 using T2w and ADC data [43]. These results highlight the intrinsic difficulty of aggressiveness classification using imaging alone but also reinforce the value of hybrid clinical-radiomic approaches.

A key strength of our approach is its methodological rigor, including the use of nested cross-validation (CV) for model development and evaluation. This strategy avoids data leakage by ensuring feature selection and hyperparameter tuning are performed within inner CV loops, while outer folds remain strictly independent for unbiased performance assessment. Such design choices increase the reliability of reported metrics and enhance model generalizability across clinical settings.

While our AUCs for ISUP (0.66), cN (0.77) and cM (0.80) may appear modest compared to some mpMRI or PET-based models, it is important to emphasize that our approach relies solely on T2w MRI and routinely available clinical data. This design prioritizes generalizability and real-world feasibility and the achieved sensitivity and balanced accuracy demonstrate clinical utility for first-line staging.

Finally, our use of SHAP-based explainability shows that radiomic features such as entropy, non-uniformity and median intensity, combined with anatomical zone indicators and PIRADSv2 scores, consistently drive predictions across all tasks. This transparency supports clinical adoption and patient-specific risk discussions.

## 1 Conclusions

This work introduces a fully automated, interpretable and resource-conscious pipeline for first-level triage in prostate cancer based on T2w MRI, radiomic features and clinical variables. The pipeline addressed three clinically relevant classification tasks: tumor aggressiveness (ISUP), LNI (cN) and distant metastasis (cM). Despite relying solely on T2w images, the models demonstrated competitive performance, particularly in the detection of metastatic disease, while maintaining high sensitivity — a key requirement for clinical screening and staging. The integration of prostate zone–specific radiomics, ComBat harmonization across centers and SHAP-based explainability contributed to both model robustness and interpretability.

## Data Availability

All data produced in the present study are available upon reasonable request to the authors

## Acknowledgments

CHAIMELEON has been funded by Horizon 2020 project (RIA, topic

DT-TDS-05-2020-AI for Health Imaging; call SC1-FA-DTS-2019-1, under Grant Agreement No. 952172).

